# Multi-modal integration of protein interactomes with genomic and molecular data discovers distinct RA endotypes

**DOI:** 10.1101/2025.07.31.25331970

**Authors:** Javad Rahimikollu, Priyamvada Guha Roy, Akash Kishore, Danica Morgan Lee, Lauren A Vanderlinden, Kiran Nazarali, Fan Zhang, Dana P Ascherman, Daniella M Schwartz, Larry Moreland, Jishnu Das

**Affiliations:** Center for Systems Immunology, Departments of Immunology and Computational & Systems Biology, University of Pittsburgh, Pittsburgh, PA, USA; Joint CMU-Pitt PhD Program in Computational Biology, Pittsburgh, PA, USA; PhD Program Department of Human Genetics, School of Public Health, Pittsburgh, PA, USA; Division of Rheumatology, University of Colorado Denver, Aurora, CO, USA; Division of Rheumatology and Clinical Immunology, Department of Medicine, University of Pittsburgh School of Medicine, Pittsburgh, PA, USA

## Abstract

**Background:** Rheumatoid arthritis (RA) is a complex autoimmune disease characterized by clinical and molecular heterogeneity, notably in the presence of anti-cyclic citrullinated peptide antibodies (CCP). CCP positivity in RA patients is associated with more severe disease progression and distinct responses to treatment compared to CCP-patients. Although previous studies have investigated cellular and molecular differences between these RA subtypes, there has been limited exploration of their genetic differences at a systems scale, taking into account underlying molecular networks.

**Methods:** Here, we use a novel multi-scale framework that couples a network-based genome-wide association study (GWAS) to functional genomic data to uncover network modules distinguishing CCP+ and CCP-RA.

**Findings:** We utilized the RACER (Rheumatoid Arthritis Comparative Effectiveness Research) cohort, comprising 555 CCP+/RF+ and 384 CCP-/RF+ RA patients, and uncovered significant differences in heritability between these two disease groups. This was followed by a network-based GWAS which uncovered 14 putative gene modules that explained genetic differences between CCP+/RF+ and CCP-/RF+ RA. Interestingly, these included many genes outside the HLA locus. Further validation through heritability partitioning and multivariate expression analyses underscored the significance of specific modules, highlighting novel genetic loci driving heterogeneity in antibody prevalence in RA. The identified modules were validated in a completely orthogonal cohort from the All of Us program. Functional analysis revealed that these modules captured critical molecular programs that not only relate to serological variation but also underlie broader functional heterogeneity in RA, including differences in synovial cell-type abundance phenotypes and variation in treatment response.

**Interpretation:** Our findings demonstrate the utility of network-based approaches in elucidating the complex genetic landscape of RA, offering new insights into the differential genetic risk factors underlying CCP+/RF+ and CCP-/RF+ RA, and paving the way for more personalized therapeutic strategies.

## Introduction

Rheumatoid arthritis (RA) is a progressive autoimmune disease that causes inflammation in the lining of synovial joints. According to the Global Burden of Diseases, Injuries, and Risk Factors Study 2021 (1), approximately 17.6 million people are affected by this chronic condition globally. In addition to clinical heterogeneity, RA is associated with substantial molecular and cellular heterogeneity. One of the primary causes of heterogeneity in RA is the presence of anti-cyclic citrullinated peptide antibodies (CCP) and rheumatoid factor (RF) in the serum. Even though the presence of these autoantibodies is a part of the diagnostic criteria for RA, patients with RA often show seronegativity for one or both (2).

The seroprevalence of CCP autoantibodies is a defining feature of heterogeneity in RA patients and is associated with differing prognoses. CCP seropositivity in RA has been associated with more aggressive disease prognosis compared to CCP seronegativity (3). Conversely, seropositivity is also associated with better responsiveness to disease-modifying anti-rheumatic drug treatment [4]. While prior work has outlined cellular and molecular differences between CCP+/RF+ and CCP-/RF+ RA (4–7), there has been limited exploration of differences in the genetic architecture of these groups. Comparisons of findings from case-control studies for CCP+/RF+ and CCP-/RF+ RA have shown that both disease endotypes have shared and distinct genetic loci underlying their pathogenic architectures (8–10). This suggests that there are differences in their genetic bases. However, this has not been systematically explored. Only one major genome-wide association study (GWAS) comparing CCP+/RF+ and CCP-/RF+ RA patients has been reported in the literature, but the findings of this study were limited to the HLA loci due to the small sample size of the cohort (11). Very few GWASs have compared genetic factors associated with both RA subtypes. Most studies have been case-control studies that considered either disease subtype but not both or have grouped them together (12–16).

A GWAS involves testing differences in allele frequency to find associations between genetic variants and phenotypes (17). While GWAS remains a cornerstone of genetic research, there are several limitations to traditional GWAS (18). One major limitation is that the statistical power for detecting associations depends on the sample size of the cohort, which implies that to identify additional disease loci beyond ones with very large effect sizes, progressively larger sample sizes are needed. This represents a major limiting factor when studying disease subsets with small cohort sizes. Given that increasing sample size to facilitate discovery of novel genetic loci is not always feasible, other methods are needed to improve discovery for smaller sample sizes. One such method is a network-based GWAS. It involves leveraging biological networks, such as protein-protein interaction (PPI) networks based on the guilt by association strategy, since interacting proteins usually are involved in the same molecular pathways and are often implicated in the pathogenesis of the same trait (19). By propagating the signal from a GWAS over the PPI network, it is possible to augment signal by borrowing strength from functionally similar genes, which were not significantly associated with the trait due to low power, and thus identify additional disease associated loci (20).

While network-based approaches have been used to study the genetics of several diseases, such as multiple sclerosis (21) and Crohn’s disease (22), they have rarely been utilized to investigate the genetic differences between two putative disease endotypes. Here, we aimed to dissect the differences in genetic architecture, beyond the HLA region, underlying CCP+/RF+ and CCP-/RF+ RA using a network-based GWAS approach. We then used heritability partitioning and functional genomic datasets to identify key network modules, including novel genetic loci beyond the HLA region, that drive heterogeneity across CCP+/RF+ and CCP-/RF+ RA. Several of these genes were replicated in an orthogonal genetic analysis using the All of Us cohort (23). Additionally, these gene modules were found to represent critical molecular processes that underlie clinical heterogeneity in RA—extending beyond autoantibody seroprevalence to include differences in synovial cell-type abundance phenotypes and variability in treatment response.

## Results

We leveraged the Rheumatoid Arthritis Comparative Effectiveness Research (RACER) cohort, which has been previously described (24), to elucidate differences at the genetic and molecular level between these subtypes of RA. Although the cohort did not intentionally recruit only Caucasian patients, the majority enrolled were Caucasian. The cohort consisted of 555 patients with CCP+/RF+ and 384 patients with CCP-/RF+ RA (Figure 1A). However, a conventional GWAS would not be sufficiently powered to elucidate differences between these disease subtypes as shown in a previous study. To overcome this limitation, we employed a network-based GWAS approach, which integrates genetic association data with prior-knowledge biological networks, such as protein interactome networks. By considering the interactions and relationships among genes, network-based GWAS can identify genetic associations that may be missed by traditional single-variant analysis (Figure 1B). Using this method, we could prioritize clusters of genes or gene modules that can shed light on functionally relevant pathways. These modules underwent further orthogonal analyses to establish significance. It is important to note that while significance was demonstrated for some, this does not preclude the possibility of significance in other modules, rather it underscores the validity of the observed signal.

**Figure 1:**
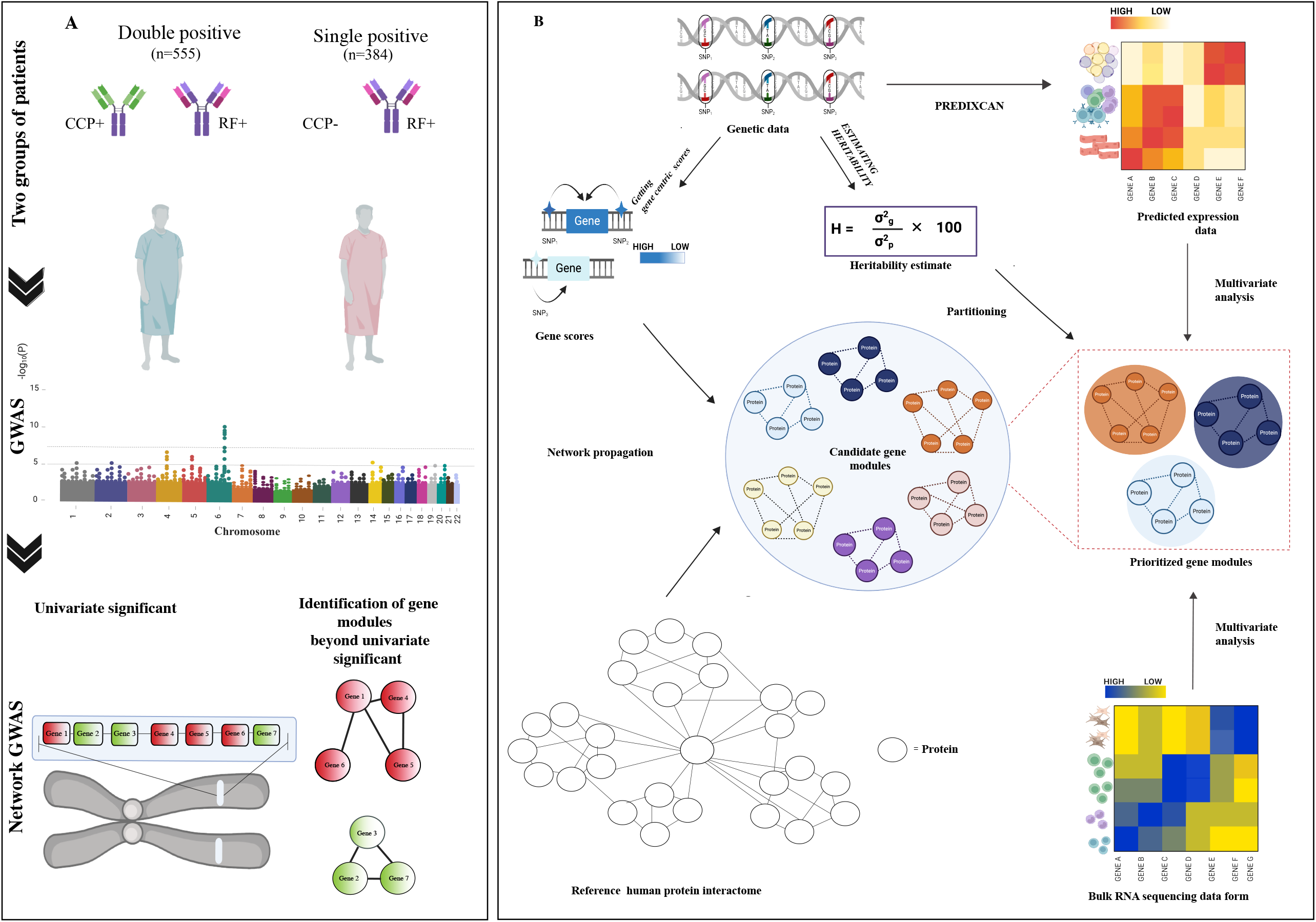
Multi-Scale Network-Informed Genetic Analyses to Identify Heritability and Phenotypic Differences Across CCP+/RF+ and CCP-/RF+ RA. A. Conceptual overview of our study B. Schematic representation of analytical pipeline used to identify and validate genetic loci underlying phenotypic differences between CCP+/RF+ and CCP-/RF+ RA

### Heritability Analysis Highlights Distinct Genetic Underpinnings of CCP+/RF+ and CCP-/RF+ RA, but Conventional GWAS Implicates only HLA Loci

To quantify differences in the genetic basis of CCP+/RF+ and CCP-/RF+ RA, we first estimated heritability, a measure of the proportion of phenotypic variation attributable to genetic factors. We computed the heritability of each subgroup using individual genotypes for the 939 subjects in the RACER cohort. After excluding variants with a minor allele frequency (MAF) below 0.05, approximately 7,200,000 single nucleotide polymorphisms (SNPs) were included in the analysis. To estimate differences in heritability between the two groups, we applied the Linkage Disequilibrium-Adjusted Kinship (LDAK) Restricted Maximum Likelihood (REML) approach (25). The cohort comprised an unequal distribution of subjects by biological sex (3.5:1 ratio of female: male subjects), consistent with higher disease incidence of RA in females. So, we accounted for sex as a fixed effect in our analysis. In addition, we designated the first principal component from principal component analysis as a fixed effect to control for any potential confounding factors arising from admixture. After accounting for these two fixed effects, the difference in heritability was estimated to be 0.31 (p-value = 0.03, Figure 2A), highlighting significant differences in the genetic factors underlying each disease subgroup. This heritability estimate implies that ∼30% of the differences in phenotype between these subgroups can be explained by genetics, and the possibility that these subgroups are indeed distinct endotypes. Our results are in line with those of a large-scale familial aggregation study conducted within a Swedish population by Frisell et al. (26), which found the heritability of CCP+/RF+ RA to be approximately 50% and that of CCP-/RF+ RA to be around 20%. These findings underscore a significant genetic difference between the groups, which is not mirrored in the findings of a previously published twin study (27).

**Figure 2:**
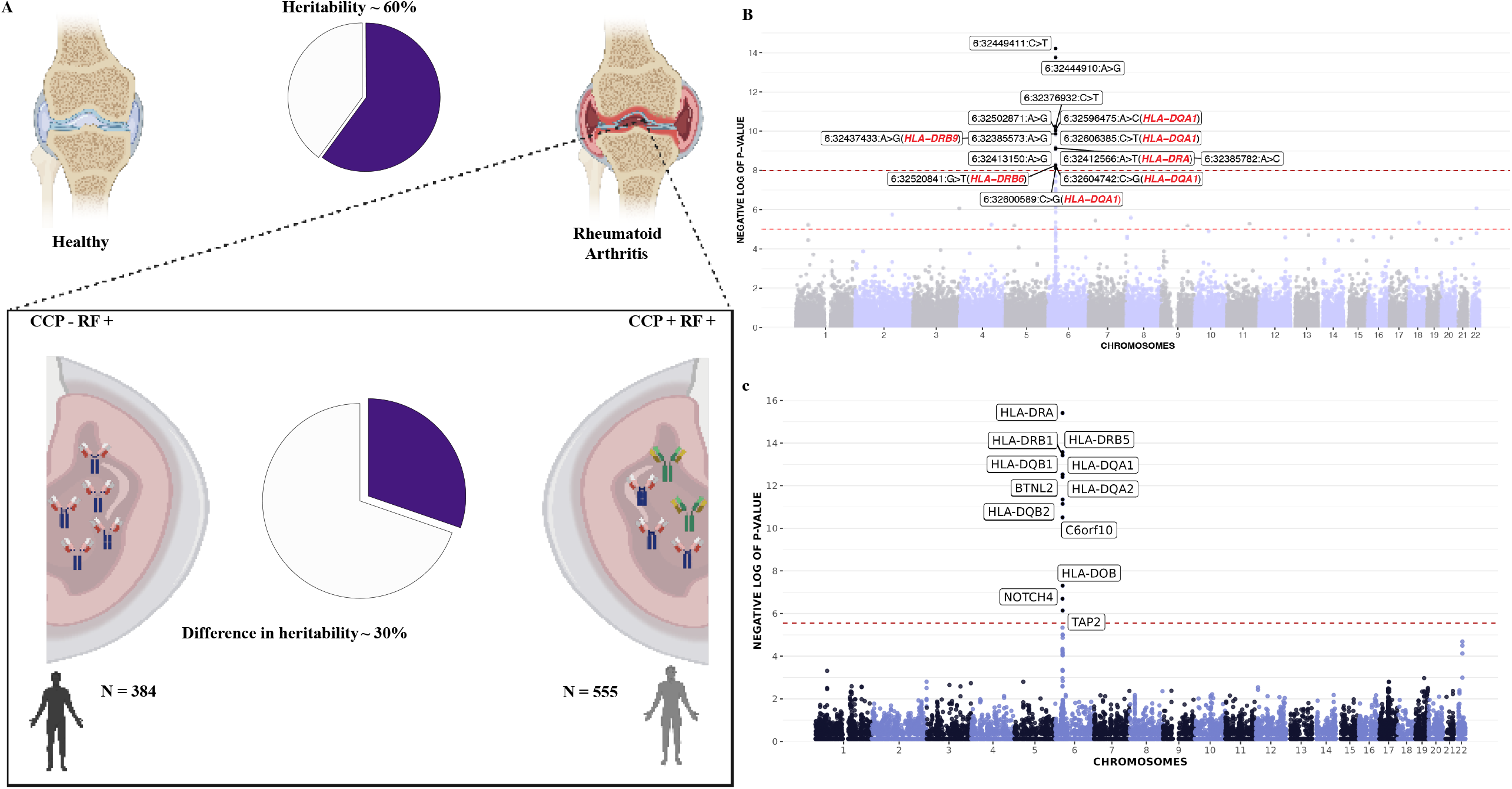
Genetic Loci Identified Using Conventional GWAS do not Explain the Difference in Heritability between CCP+/RF+ and CCP-/RF+ RA. A. Difference in heritability between CCP+/RF+ and CCP-/RF+ RA within the RACER cohort B. Manhattan plot for GWAS comparing CCP+/RF+ and CCP-/RF+ RA where SNPs with significant association statistics are labeled C. Manhattan plot for gene-based association analyses comparing CCP+/RF+ and CCP-/RF+ RA where genes with significant association statistics are labeled

While the earlier heritability analyses provide insights into how much of the phenotypic differences can be attributable to genetic factors, it does not highlight which genetic loci underlie these differences. To identify these genetic factors, we conducted a GWAS, adjusting for the first principal component of genotype and sex. Our analysis identified 2441 genetic variants with association p-values that met the Bonferroni-corrected significance threshold of 5 × 10^−8^. All the genome-wide significant variants were confined to the HLA region on chromosome 6, and a handful of additional variants surpassed a nominal p-value threshold of 5 × 10^−5^ (Figure 2B). While these findings were in line with those reported in the previous GWAS comparing CCP+/RF+ and CCP-/RF+ RA cases (11), there were clearly a large number of missed genetic associations. When the difference in heritability across the two groups was estimated using only the significant SNPs identified in the GWAS, it was found to be ∼3%, i.e., only 10% of the overall 30% estimated difference in heritability across CCP+/RF+ and CCP-/RF+ individuals was explained by the significant GWAS SNPs. This suggests that due to stringent false discovery rate control, there were several false negatives, i.e., missed associations (Figure 2B). Further, we performed univariate gene association tests using LDAK, identifying significant associations with several genes, such as *HLA-DRB1, HLA-DRB5*, and *HLA-DRA*, all within the HLA region (Figure 2C). While this gene-based approach extended beyond locus-level associations, it still predominantly highlighted only significant associations within the HLA region. However, it is essential to recognize that univariate gene set tests inherently analyze each gene individually and are underpowered as they do not consider the interrelationships or combined effects between multiple genes.

### Network GWAS Unveils Additional Loci Underlying the Phenotypic Differences between CCP+/RF+ and CCP-/RF+ RA

With this cohort size, a conventional GWAS is underpowered to detect associations beyond the HLA locus. To address this, we adopted an alternate strategy motivated by network-based GWAS. Network-based GWAS is an approach that integrates genetic association data with biological networks to identify genes implicated for a given phenotype. High-scoring genes that are randomly distributed across the network may not reflect meaningful biological associations. In contrast, moderately scoring genes that are highly proximal within the network are more likely to indicate a real signal, as they often participate in the same pathways and contribute to the same traits. Additionally, even when the significance level is below the stringent Bonferroni corrected threshold, the presence of a single high-scoring gene can still indicate a significant finding. This is because proteins that interact usually operate in similar molecular pathways and are commonly linked to the same trait (20).

In our analysis, we utilized the LDAK REML method to estimate a comprehensive gene-centric score, which accounted for not only coding variants but also non-coding variants in *cis*-regulatory elements (i.e. enhancers) that modulate gene expression (Figure 3A). The score summarized the combined impact of these variants into gene-centric scores taking into account the underlying linkage disequilibrium structure. We then propagated these scores over a high-quality reference protein interactome (28) using a random-walk-with-restart algorithm implemented in HotNet2 (29). The network propagation algorithm converged on high-scoring modules where the scores corresponding to the individual genes encoding the proteins did not necessarily meet the univariate significance threshold, but the module met an overall false discovery threshold of 0.1 using permutation testing (network edge swaps). The use of permutation testing is a stringent way to ensure that we converge on these modules only using the true protein network topology and structure and not shuffled networks that are otherwise perfectly matched in terms of degree distribution. Using this approach, we identified 14 statistically significant network modules (Supplementary Table 1). These modules comprised 193 genes (Figure 3B), including genes from the HLA region and a wide array of other molecular pathways, which have not been previously implicated in the phenotypic differences between CCP+/RF+ and CCP-/RF+ RA.

**Figure 3:**
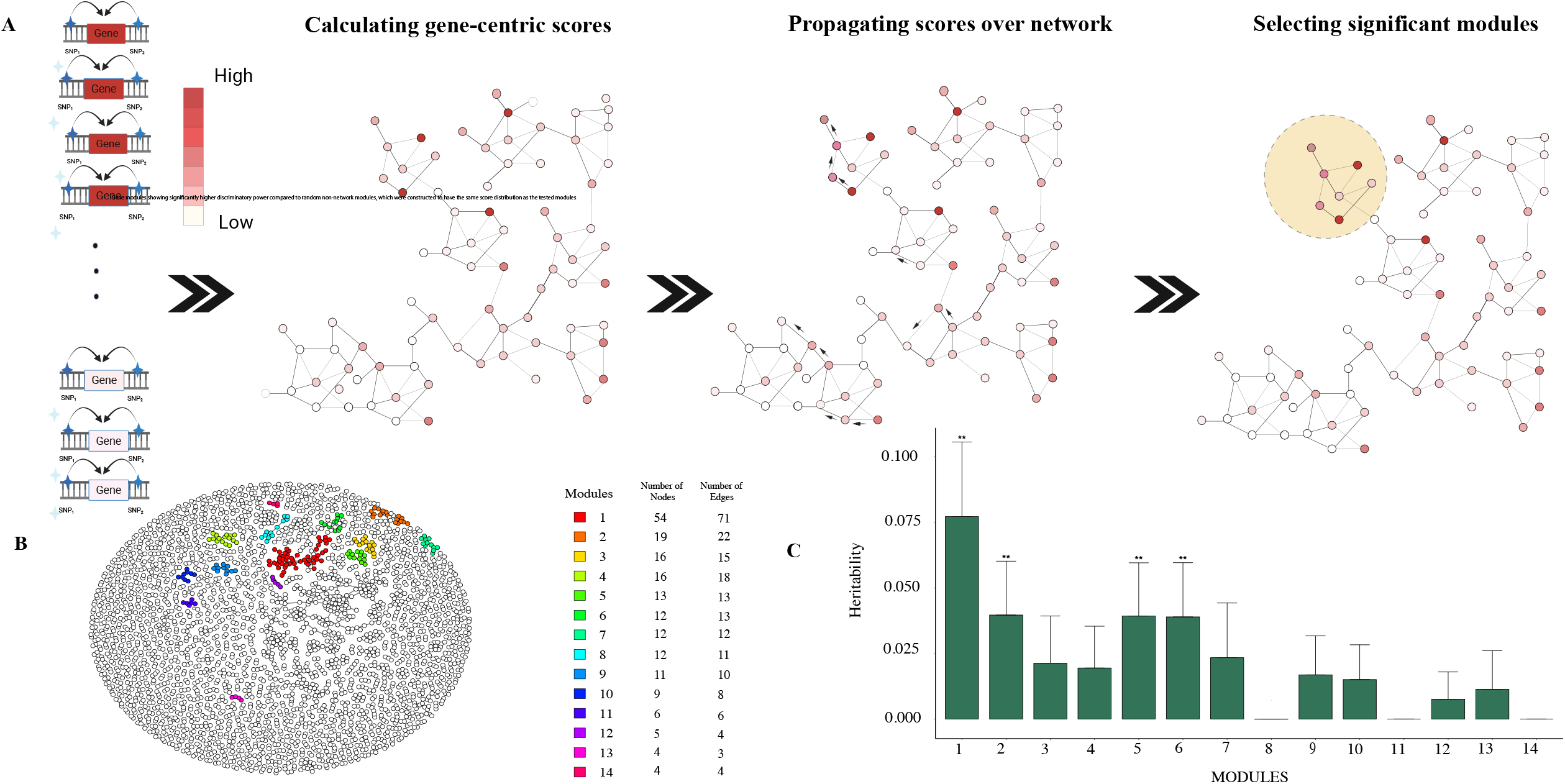
Network GWAS Identifies Additional Associations Underlying Genotypic Differences between CCP+/RF+ and CCP-/RF+ RA. A. Schematic explaining how network propagation was used to identify genetic loci underlying the phenotypic differences between CCP+/RF+ and CCP-/RF+ RA B. Network-based GWAS analysis identifies 14 significant modules as candidates explaining genotypic and heritability differences between CCP+/RF+ and CCP-/RF+ RA C. Bar plot showing per module contribution to the difference in heritability between CCP+/RF+ and CCP-/RF+ RA, where ** represent significant contributions as measured by the LDAK REML p-value

Though we employed stringent false discovery rate control, our analyses identify modules that correlate with the difference in phenotype seen between CCP+/RF+ and CCP-/RF+ RA. To move beyond correlations, we interrogated these modules in terms of their ability to explain heritability partitioning as well as against several functional genomic datasets. This multi-step prioritization scheme helps identify the functional significance of the modules using multiple orthogonal analyses and datasets. The use of a combination of multiple layers of evidence (genetic, cellular, and molecular) from different datasets helps us converge on the most robust and biologically meaningful modules that explain heterogeneity between these two subtypes of RA.

### Candidate Gene Modules Explain the Entire Difference in Heritability Estimate

To better understand the relative importance of each module, we estimated the contribution of each module to the total heritability estimate for the traits. Given that certain parts of the genome contribute disproportionately to heritability, we hypothesized that functionally important gene modules will have higher heritability contributions. Significant SNPs identified from the conventional GWAS only identified the HLA locus and accounted for only a small proportion (∼3%, i.e., 10% of the estimated difference of ∼30%) of the estimated difference in heritability between the 2 subgroups. We hypothesized that SNPs mapped to the significant modules identified above could account for the missing heritability. We estimated heritability for each candidate gene module using the LDAK REML approach. Combining heritability across all network modules resulted in an estimate of a ∼31% difference in heritability across the two disease subtypes, which was comparable to the estimate obtained using all SNPs in the set. Moreover, partitioning heritability among the modules can help us prioritize the modules based on their genetic contribution to the overall heritability. Interestingly, in addition to the HLA module (module 1) demonstrating a statistically significant heritability of 0.075, modules 2, 5, and 6 by themselves also showed significant contributions to the overall difference heritability across the two disease subgroups (Figure 3C). These modules were functionally interpreted using GeneSetAI, a machine learning tool that assigns biologically meaningful labels based on gene content and known functional annotations (30).

### Multivariate Analysis using Sorted Bulk RNA-seq Data Reveals Cell Type Specific Signals for Candidate Gene Modules

In addition to explaining heritability, we hypothesized that functionally important network modules would correspond to genes whose expression programs are significantly different between doubly seropositive (CCP+/RF+) RA and singly seropositive RA. So, we performed an orthogonal validation analysis using a dataset of bulk RNA-seq from the Accelerating Medicines Partnership Rheumatoid Arthritis consortium (AMP RA) phase I data (31). Interestingly, while this cohort included CCP+/ RF+ RA patients, most of the singly seropositive patients were CCP+/RF- and a few were CCP-/RF+. However, in the RACER cohort, the singly seropositive patients were CCP-/RF+. Despite this difference, the analysis allows us to evaluate whether the identified modules are overall discriminatory between doubly and singly seropositive RA.

We tested whether the expression of genes encoding proteins within the identified network modules could significantly discriminate CCP+/RF+ RA from other subtypes across different cell types, using the area under the Receiver Operating Characteristic (ROC) curve (AUC) as the performance metric. We performed permutation testing by randomly selecting size-matched gene sets from all genes in the bulk RNA-seq data. In fibroblasts, the interferon signaling and extracellular matrix remodeling module (module 1) was discriminatory between doubly vs singly seropositive RA (Figure 4A). Genes coding for proteins comprising the immune response and inflammation modulation and complement system activation and regulation modules (modules 4 and 5, respectively) were found to be discriminatory in T cells (Figure 4B). The cytokine signaling and immune response modulation module (module 7) was significant in B cells (Figure 4C), and the interferon signaling and extracellular matrix remodeling, TGF-beta/BMP signaling in developmental processes, protein folding and glycosylation in the endoplasmic reticulum, and chemokine-mediated signaling in immune response modules (modules 1, 2, 10, and 13, respectively) were significant in monocytes (Figure 4D).

**Figure 4:**
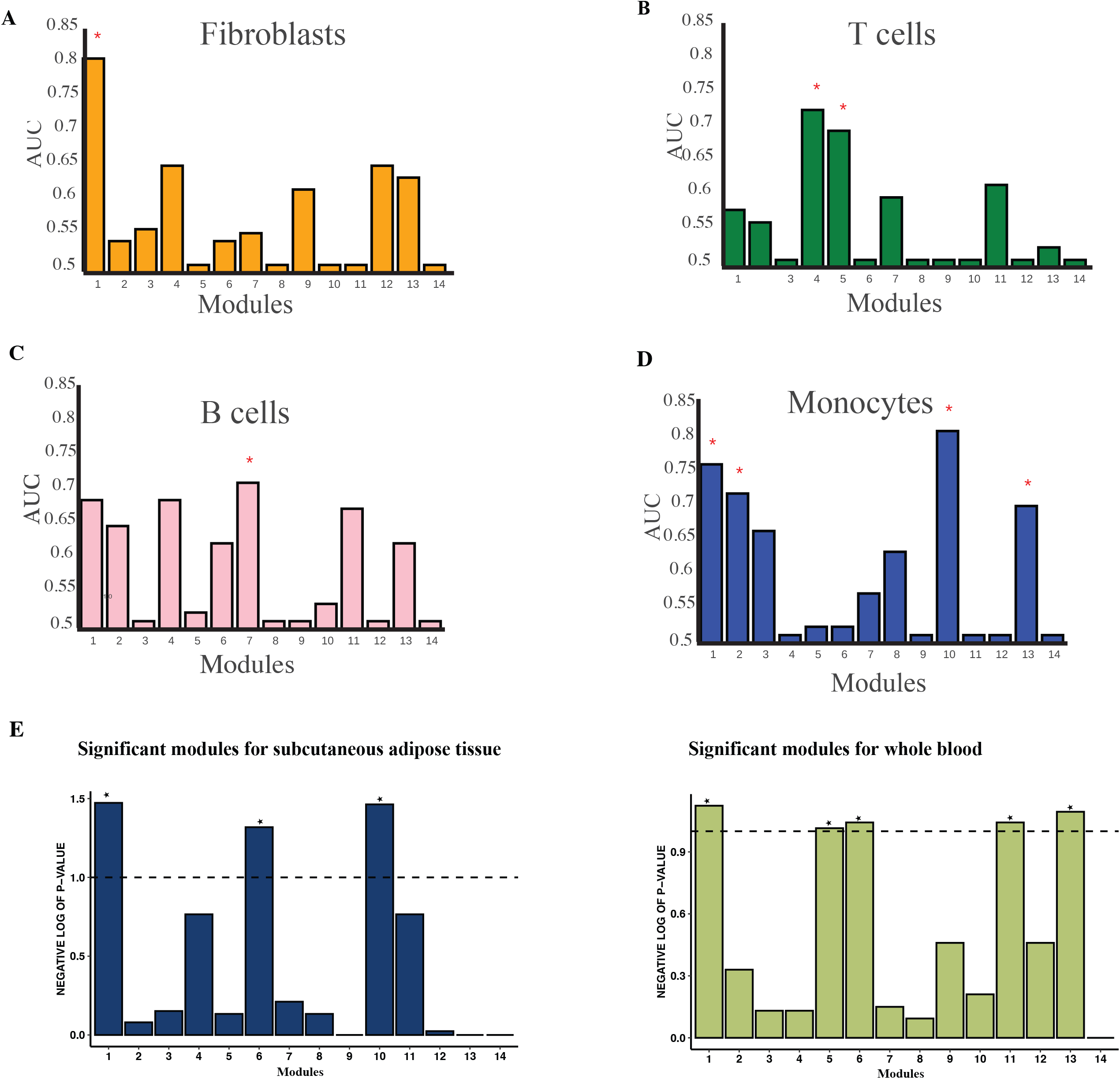
Multivariate Analyses of Sorted Bulk RNA-seq Data and Imputed Gene Expression Data Reveal Cell Type and Tissue Specificity of Candidate Gene Modules. A. Bar plot showing the discriminative ability of a multivariate logistic regression model based on gene expression profiles for each module in fibroblasts, where AUC values represent the model’s ability to discriminate CCP+/RF+ RA from other RA subsets and ‘*’ represent the model’s performance against a score distribution matched module B. Bar plot showing the discriminative ability of a multivariate logistic regression model based on gene expression profiles for each module in T cells, where AUC values represent the model’s ability to discriminate CCP+/RF+ RA from other RA subsets and ‘*’ represent the model’s performance against a score distribution matched module C. Bar plot showing the discriminative ability of a multivariate logistic regression model based on gene expression profiles for each module in B cells, where AUC values represent the model’s ability to discriminate CCP+/RF+ RA from other RA subsets and ‘*’ represent the model’s performance against a score distribution matched module D. Bar plot showing the discriminative ability of a multivariate logistic regression model based on gene expression profiles for each module in monocytes, where AUC values represent the model’s ability to discriminate CCP+/RF+ RA from other RA subsets and ‘*’ represent the model’s performance against a score distribution matched module E. Bar plot showing the discriminative ability of a multivariate logistic regression model based on predicted gene expression for each module in subcutaneous adipose tissue, where -log(p-values) represent the discriminatory power of the modules and ‘*’ represent models with - log(p-value) ≥ 1 F. Bar plot showing the discriminative ability of a multivariate logistic regression model based on predicted gene expression for each module in whole blood, where -log(p-values) represent the discriminatory power of the modules and ‘*’ represent models with -log(p-value) ≥ 1

### Multivariate Analysis using Imputed Expression from RACER Cohort Prioritizes Specific Candidate Gene Modules

To further validate our findings at the expression level, we imputed the expression levels of genes in the 14 modules for subjects in the RACER cohort based on their individual genotypes. We utilized PrediXcan (32), a regularized regression approach that leverages individual genetic variation to predict corresponding expression levels for each subject. This is an orthogonal but equally important approach as we used imputed matched (at a per-subject level) gene expression in this analysis as opposed to an orthogonal cohort in the prior analysis. By using the PrediXcan model, we were able to impute expression levels for both adipose tissue and whole blood. Consistent with the prior transcriptomic analysis using data from the RACER cohort, this assessment revealed that several of the 14 identified modules were again significantly discriminatory between CCP+/RF+ and CCP-/RF+ RA in adipose tissue and whole blood (Figure 4E).

### Network GWAS Identifies Cell Type Specific Modules That Stratify Disease Endotypes

Our study introduced a robust and innovative method for identifying genetic factors underlying the phenotypic difference between CCP+/RF+ and CCP-/RF+ RA patients. Utilizing a network-based GWAS approach, we identified 14 candidate gene modules (Figure 5A). Five of these modules: *interferon* signaling and extracellular matrix remodeling (module 1, Figure 5B), complement system activation and regulation (module 5, Figure 5C), TGF-beta/BMP signaling in developmental processes (module 2, Figure 5D), protein folding and glycosylation in the endoplasmic reticulum (module 10, Figure 5D) and protein processing and cellular transport (Supplementary Figure 1) modules successfully passed two out of the three stringent validation analyses. Together, these modules comprise 107 genes. The largest module, comprising 54 genes, included genes from within the HLA region and other immune signaling genes. But the other modules encompassed a wide range of biological processes. Despite the vast search space of > 17,000 proteins in the entire network, our method demonstrates a high level of specificity by prioritizing 107 proteins within the modules (Figure 5A). Interestingly, these modules contain genes with both high and low gene scores, highlighting the potential of our approach to identify genes that may be overlooked in traditional GWAS analyses.

**Figure 5:**
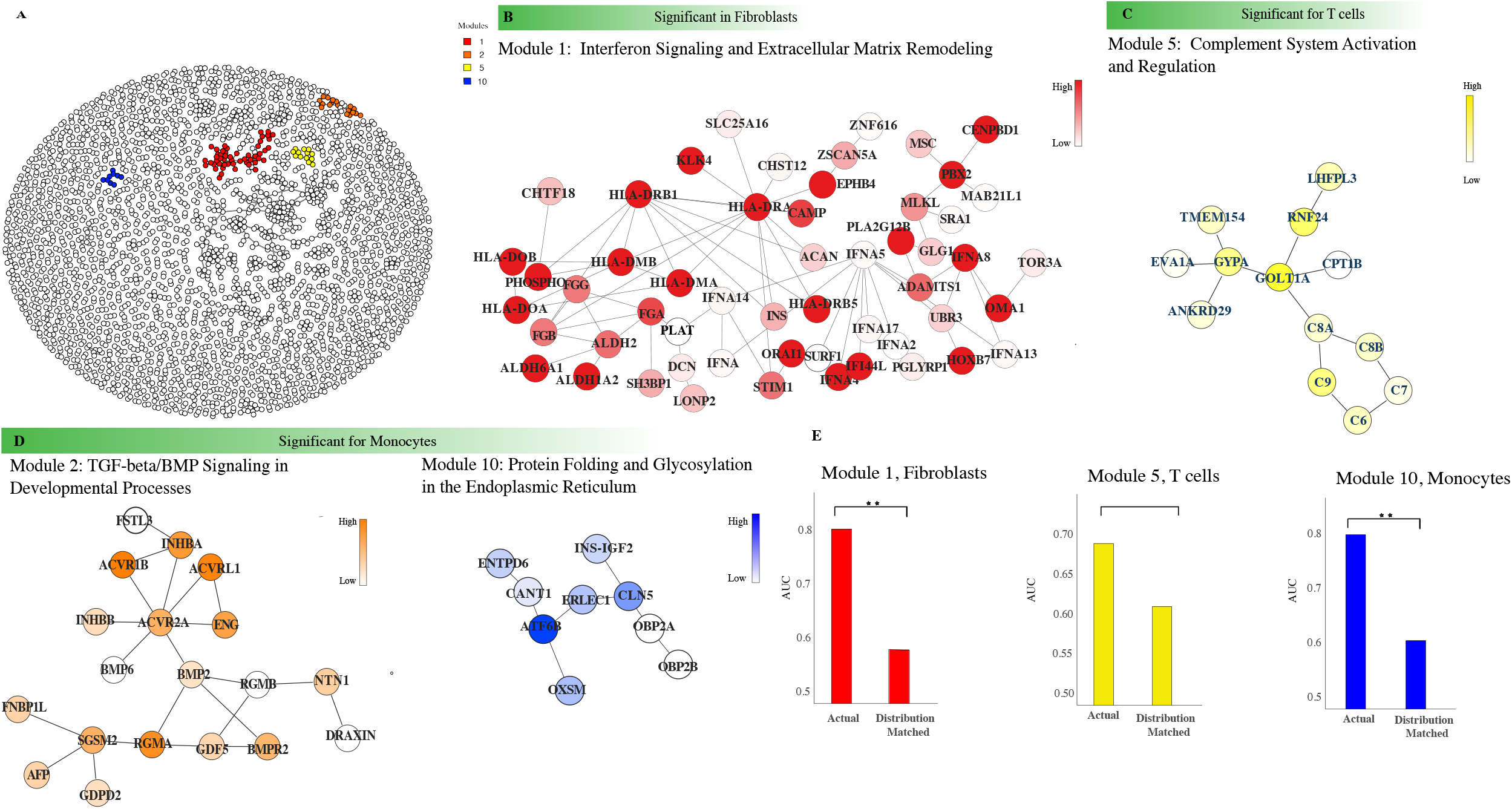
Prioritized Gene Modules Reveal Novel Cross-talk between HLA-Type 1 IFN underlies the Phenotypic Differences Seen between CCP+/RF+ and other RA subsets. A. Visualization of prioritized gene modules in the overall network (prioritized modules are colored) B. Module with significant performance in multivariate analysis using expression data from fibroblasts C. Module with significant performance in multivariate analysis using expression data from T cells D. Module with significant performance in multivariate analysis using expression data from monocytes E. Gene modules showing significantly higher discriminatory power compared to random distribution matched gene sets

Our analysis identified a significant module comprising complement system genes, including *C9, C8A, C8B, C7*, and *C6* (Figure 5C). The complement system is a crucial part of the innate immune response, and its dysregulation has been implicated in autoimmune diseases such as RA. Notably, the presence of *C9, C8A, C8B, C7*, and *C6*, genes encoding components of the membrane attack complex, in module 5 (complement system activation and regulation) indicates a potential involvement of complement-mediated cytotoxicity and subsequent tissue damage mechanisms in RA (Figure 5C). Overall, our findings shed new light on the complex interplay of genes and proteins in the mechanism of RA and open new avenues for further research and potential therapeutic interventions.

Gene modules represent structured sub-networks rather than merely groups of unrelated genes. To highlight the importance of this connectivity structure within our identified modules, we compared the performance of candidate gene modules against ‘distribution-matched’ control modules. These control modules were constructed by randomly sampling genes according to the distribution of node heat scores in each candidate module, deliberately disregarding the underlying protein interaction network structure. Node heat scores were categorized into three bins—high, medium, and low—to ensure the number of genes selected in each bin of the control modules matched those in the corresponding candidate modules. For example, if a gene module contained four high and one medium heat score genes, an equal number of genes were randomly selected from the respective heat bins to construct the control module. This comparison allowed us to specifically measure the contribution of the PPI network structure to the candidate modules, allowing us to assess whether the significant associations identified by the network-based method were due to the underlying biological relationships between genes, rather than random chance (Figure 5E). Even under this stringent control, we observed significant differences in the predictive power of our modules compared to the ‘distribution-matched size-matched’ controls. Specifically, these differences were notable for the interferon signaling and extracellular matrix remodeling module (module 1) in T cells, and the protein folding and glycosylation in the endoplasmic reticulum module (module 10) in monocytes. Together, these results indicate that the identified modules underlie key phenotypic differences in doubly vs singly seropositive RA.

### Validation of Selected Gene Modules Using Orthogonal Genetic Data from the All of Us Cohort

To independently validate the gene modules identified from network GWAS of the RACER cohort, we leveraged genotype and phenotype data from the All of Us cohort (23). We restricted our analysis to European ancestry individuals who were positive for RF (RF > 20 IU/mL), yielding a cohort of 248 RA patients—a population composition like that of the RACER cohort. These samples were further stratified based on anti-CCP antibody status into double-positive (CCP+/RF+; *n* = 93) and single-positive (CCP-/RF+; *n* = 155) subgroups, with CCP values greater than 20 IU/mL considered positive (Figure 6A). A GWAS comparing these two groups revealed significant enrichment at the HLA class II locus on chromosome 6 (Figure 6B), consistent with findings in the RACER cohort (Figure 2B). We used LDAK to aggregate SNP scores for each gene and compared the scores of genes from our modules to distributions derived from random, size-matched gene sets. Notably, two modules – interferon signaling and extracellular matrix remodeling (module 1) and complement system activation and regulation (module 5) showed significantly higher genetic scores relative to random expectations (Figure 6C), highlighting the robustness of our findings and the importance of these genes in shaping the phenotypic differences between the endotypes.

**Figure 6:**
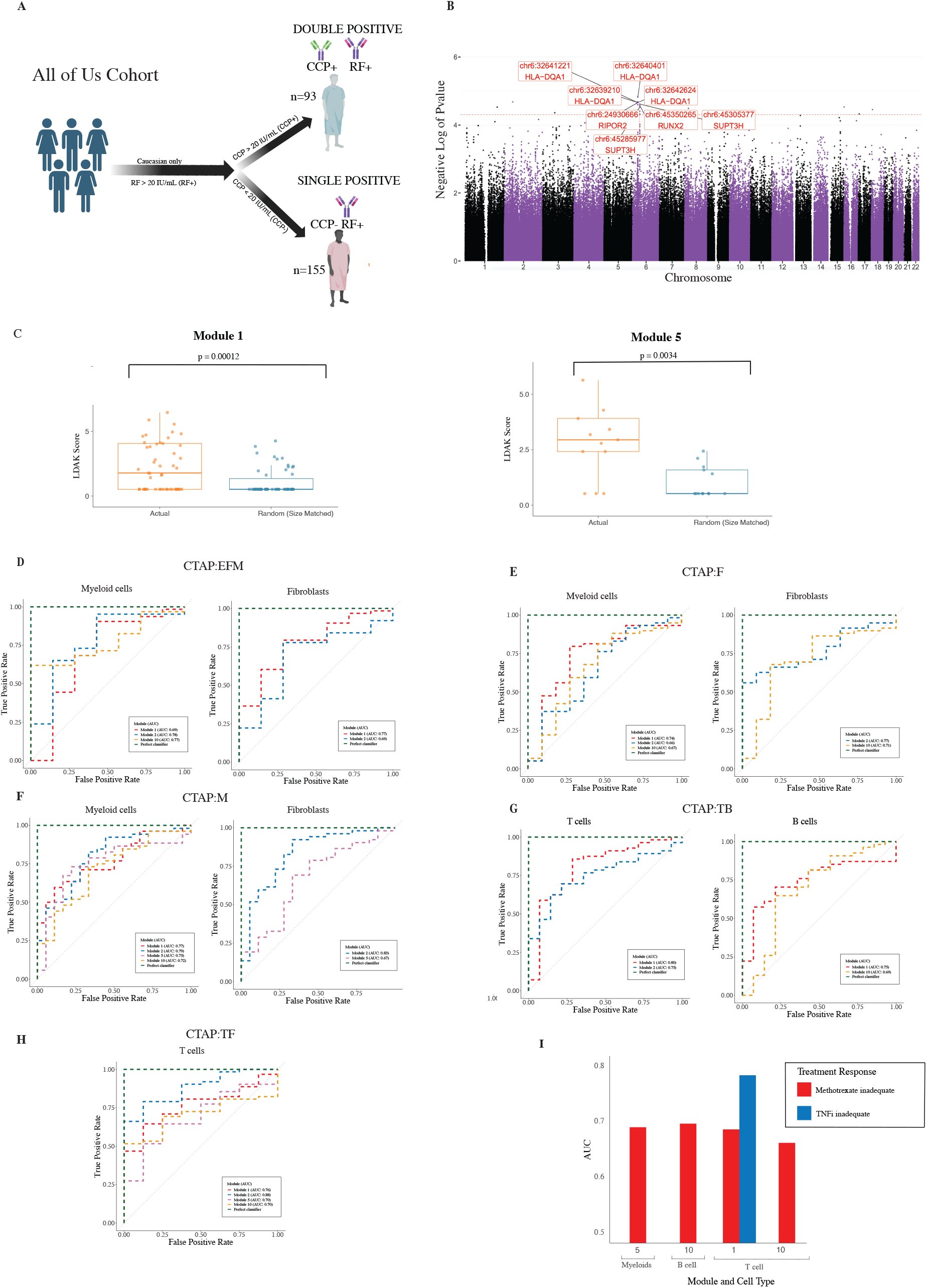
Functional Validation of CCP-Associated Gene Modules in Predicting Synovial CTAP using scRNA-seq and ROC Analysis. A. Schematic of All of Us cohort selection and serological stratification Participants from the All of Us cohort were filtered to include only Caucasian individuals who were RF+ (RF > 20 IU/mL). These individuals were further stratified based on CCP antibody levels into two serological RA endotypes: CCP+/RF+ (double positive; n = 93) and CCP−/RF+ (single positive; n = 155) B. Manhattan plot for GWAS comparing CCP+/RF+ and CCP-/RF+ RA in All of Us cohort where SNPs with significant association statistics are labeled C. Comparison of LDAK scores for prioritized gene modules from network GWAS versus size-matched random gene sets in the All of Us cohort D. ROC curves evaluating the ability of gene modules to distinguish the CTAP-EFM synovial phenotype from other CTAP phenotypes using scRNA-seq data E. ROC curves evaluating the ability of gene modules to distinguish the CTAP-F synovial phenotype from other CTAP phenotypes using scRNA-seq data F. ROC curves evaluating the ability of gene modules to distinguish the CTAP-M synovial phenotype from other CTAP phenotypes using scRNA-seq data G. ROC curves evaluating the ability of gene modules to distinguish the CTAP-TB synovial phenotype from other CTAP phenotypes using scRNA-seq data H. ROC curves evaluating the ability of gene modules to distinguish the CTAP-TF synovial phenotype from other CTAP phenotypes using scRNA-seq data I. Selected gene modules are discriminative between different treatment responses

### Prioritized Gene Modules Capture Molecular Processes that Explain Heterogeneity in RA beyond Seroprevalence of Autoantibodies

Beyond serotype-based heterogeneity, RA patients also display additional clinical variation, one aspect of which is captured by cell-type abundance phenotypes (CTAPs). CTAPs reflect distinct synovial inflammatory microenvironments, ranging from lymphocyte-rich (CTAP-TB, CTAP-TF) to myeloid-or fibroblast-dominated profiles (CTAP-M, CTAP-F, CTAP-EFM) and have been shown to predict differential treatment responses. We next sought to test whether the gene modules identified through network GWAS as distinguishing CCP+/RF+ and CCP-/RF+ RA endotypes also contribute to the phenotypic differences between CTAPs. To do this, we performed functional validation using single-cell RNA-seq (scRNA-seq) data from sorted synovial cell populations. We generated ROC curves to quantify the discriminatory power of each gene module in distinguishing individual CTAPs (Figures 6D-H), with AUC values indicating stronger predictive performance.

CTAP□EFM, characterized by an enrichment of endothelial cells, fibroblasts, and myeloid cells, is associated with a comparatively less inflammatory microenvironment than the lymphocyte□rich CTAP subtypes and reflects a transitional or remodeling state of the synovium (33). Of our candidate gene modules, gene expression models for the TGF-beta/BMP signaling in developmental processes (module 2) and interferon signaling and extracellular matrix remodeling modules (module 1) showed the best discriminatory performance for CTAP□EFM (Figure 6D). These modules were also able to discriminate CTAP-F, which shows an enrichment for fibroblasts, from other CTAPs (Figure 6E). The ability of these modules to distinguish CTAPs that are both stromal-enriched and characterized by lower levels of inflammation underscores the functional relevance of the underlying molecular programs captured by these modules. These gene modules represent key processes associated with inflammatory microenvironments and ECM remodeling, highlighting their relevance not only to seroprevalence of autoantibodies but also to broader clinical heterogeneity observed in RA. CTAP-M, the myeloid-enriched cell-type abundance phenotype, is characterized by a high abundance of monocytes and macrophages, with relatively low infiltration of lymphocytes. All four gene modules showed good discriminatory performance for this phenotype in myeloid cells (Figure 6F). Notably, CTAP-M was found to be associated with CCP-negative status (33). While the modules do capture underlying differences in serological status, their strong discriminatory capacity in CTAP-M suggests that they extend beyond serostatus alone. Their performance likely reflects sensitivity to additional biological processes, such as myeloid-driven inflammation, that define this phenotype and are not solely explained by autoantibody levels.

CTAP-TB and CTAP-TF represent lymphocyte-enriched phenotypes, marked by high infiltration of lymphocytes within the synovium. Both these phenotypes were more commonly seen in patients with CCP positive synovium, which has been associated with higher lymphocyte infiltration compared to CCP negative synovium (33). Both CTAP-TB and CTAP-TF exhibit transcriptional signatures of adaptive immune activation, including elevated expression of T follicular helper cell markers, immunoglobulin genes, and cytokines such as IL-21, consistent with the presence of functionally active T–B cell interactions. Notably, the interferon signaling and extracellular matrix remodeling module (module 1) exhibited strong discriminatory power for these CTAPs (Figure 6G and H), highlighting the role of these gene modules in adaptive immune–driven inflammation. Additionally, the good performance of the TGF-β/BMP signaling in developmental processes module (module 2) across the two phenotypes suggests that TGF-β signaling may contribute to the regulation of lymphocyte activity and potentially influence B cell activation and autoantibody production in these CTAPs.

Further, predicting patient response to specific therapeutic regimens remains a significant clinical challenge in RA, given considerable clinical and molecular heterogeneity of the disease. We investigated whether cell type specific gene modules from our network-based GWAS could discriminate between patients showing inadequate responses to methotrexate or TNF inhibitors (TNFi) (Figure 6I). Indeed, the identified gene modules were discriminatory between different treatment regimens for various cell types. The T cell-associated gene module 1 was significantly predictive of inadequate TNFi responses. Moreover, the modules linked to Myeloid cells, B cells, and T cells displayed a predictive capability for methotrexate inadequacy (Figure 6I). Together, these suggest that the identified modules are functionally relevant and capture clinically relevant heterogeneity in RA beyond the seroprevalence of specific autoantibodies.

## Discussion

Differences in seroprevalence for autoantibodies like CCP represent a primary feature of clinical heterogeneity seen in RA. However, this heterogeneity is not just phenotypic but represents a spectrum of disease states. Several studies have shown that CCP+/RF+ and CCP-/RF+ RA are driven by distinct genetic architectures, but most of these studies were conducted using only CCP+/RF+ or CCP- /RF+ RA samples (8–10). Only one GWAS comparing these two endotypes directly has been conducted (11), which reported significant associations only within the HLA loci. While the HLA loci is an important contributor to the phenotypic differences seen between the two endotypes, it cannot sufficiently explain the corresponding differences in heritability. Therefore, we employed a network-based GWAS approach to discover other loci that could explain the differences in phenotype between the two disease endotypes.

GWAS remains the predominant method for identifying genetic loci associated with any trait. Genetic analysis using only CCP+ or CCP-RA have identified both common and independent loci for each disease. These include loci both inside and outside the HLA region. *ANKRD55, STAT4, C5orf30, PTPN22, ELMO1, RUNX1* and *BLK* have been associated with both disease subsets (34–36). Within the HLA region, HLA-DR4 is associated with CCP+/RF+ RA (37) and HLA-DR3 with CCP-/RF+ RA (38). Most of these associations have been obtained by case-control studies, and till date only one case-case GWAS between CCP+/RF+ and CCP-/RF+ RA has been reported (11). However, due to limited sample size, this study could not identify loci beyond the HLA region.

Biological networks, such as PPI networks, can be leveraged in conjunction with GWASs to discover key genetic associations with limited sample sizes. Given that interacting genes are involved in common molecular pathways, the concept of guilt by association can be used to help prioritize disease associated genes. It is possible to consider all direct interactors of disease associated genes as putatively causal, but this would result in a high false positive rate, and it would also miss important genes that are connected to multiple disease genes through longer path lengths (20). Network propagation of association signals can overcome this limitation, as it addresses the issue by down-weighting potentially erroneous predictions supported by a single path and promoting true causal genes, which may be missed despite being well-connected to the prior list (20). We estimated gene level association statistics for our analysis between CCP+/RF+ and CCP-/RF+ RA patients and propagated these signals across the PPI network to identify gene modules that might explain the phenotypic differences seen between the two endotypes. Using stringent false discovery rate control, we identified 14 network modules that putatively explain differences between the two RA endotypes. We further performed validation analyses using heritability partitioning and functional genomic datasets to prioritize 5 modules. Genes from these modules were replicated in an orthogonal genetic cohort. Moreover, these prioritized modules were found to capture other forms of clinical heterogeneity seen in RA, such as CTAPs and variation in treatment response.

Our study provides several key novel insights. Through our analyses, we were able to discover genes that were not previously associated with the difference seen between CCP+ and CCP-RA. We show that network propagation is a valuable tool for amplifying association signals for small sample sizes. The techniques used in this study are broadly applicable across contexts and can be used to solve key issues that conventional GWASs suffer from including missing heritability and many false negatives.

The prioritized gene modules recapitulate well-established mechanisms in RA pathogenesis while also providing novel insights into disease heterogeneity. Previous studies have shown that excessive complement activity can lead to the destruction of healthy tissue, exacerbating inflammatory processes central to RA (40). This is particularly relevant in CCP+ RA, where the presence of autoantibodies can result in formation of immune complexes that can activate the complement response (41). This could explain the more aggressive phenotype for CCP+ RA compared to CCP-RA. Understanding the role of complement system activation in RA, particularly in the context of complement system activation and regulation module (module 5), could open new avenues for therapeutic intervention. Targeting the components of the membrane attack complex or modulating its activity may offer a strategy to mitigate tissue damage and inflammation in RA, particularly in those with CCP+ disease.

Our study also has certain limitations. Our analysis included only patients of European ancestry. Therefore, the findings of this study may not be transferable to other populations. Nevertheless, this is one of the first studies to use network propagation in conjunction with case-case GWAS to identify gene modules underlying the phenotypic differences between CCP+/RF+ and CCP-/RF+ RA.

## Methods

### Cohort Description

In this study, we used data from the University of Pittsburgh RACER registry, which consisted of prospectively enrolled RA patients recruited through the University of Pittsburgh Medical Center (24). The cohort included 555 patients with CCP+/RF+ RA and 384 CCP-/RF+ RA. Genotype data was collected for all 939 patients using MegaChip. We filtered subjects based on the quality of raw genotypes as well as relatedness to exclude highly related subjects (kinship value of greater than 0.12 - same family) as they are likely to introduce biases. A total of 7,173,418 SNPs were imputed using the Sanger imputation server (42). This study was approved by the University of Pittsburgh Institutional Review Board (protocol 19090282) and adhered to the principles outlined in the Declaration of Helsinki.

### Genome Wide Association Analyses Adjusting for Appropriate Covariates

Phenotypic data for each subject was represented by a vector ‘Y’ with 939 entries, corresponding to the phenotypic values 1 for CCP+/RF+ and 0 for CCP-/RF+ RA. Genotype data for each individual across the 7,173,418 SNPs of the gene of interest was obtained and stored in a 939 by 7,173,418 genotype matrix ‘X’. Each row of ‘X’ represented an individual’s genotype data, and each column corresponded to a specific SNP.

To account for potential confounding factors, a matrix of covariates, ‘W’, with dimensions 939 by 2, was included in the analysis. These covariates were biological sex and the first principal component derived from the principal component analysis of the genotype data. Quality control (QC) was conducted using PLINK to ensure the integrity and reliability of the genotype data. Specifically, we applied a genotype missingness filter of 1% (−-geno 0.01) to exclude SNPs with a high proportion of missing data and an individual missingness filter of 1% (−-mind 0.01) to remove samples with excessive missing genotypes. Additionally, we implemented a MAF threshold of 5% (−-maf 0.05) to exclude rare variants.

### Heritability Estimation using LDAK

We used the LDAK model to estimate heritability of the phenotypic trait. To determine the difference in heritability, we began by computing the genetic relatedness matrix (GRM) using the ldak5.2 linux executable obtained from the LDAK website. After obtaining the GRM, we applied the –reml option to calculate heritability differences (43).

Unlike the uniform model, which assumes equal heritability contributions from each SNP, the LDAK model assigns weights to the expected heritability of each SNP proportional to ***q***_***j***_, as presented in Equations (1,2):

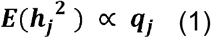

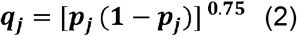

Where 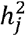 Represents the heritability of SNP j and *q*_*j*_ denotes the SNP-specific weight 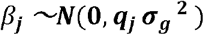 The heritability model incorporates both fixed effects (θ) and random effects **(β**). In our study, sex and the first principal were considered as fixed effects, while SNP effects 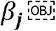 were treated as random effects. The model defines heritability as the proportion of phenotypic variance explained by the genetic variance:

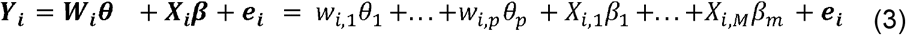

where *h*^2^ represents the heritability, 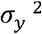 phenotypic variance, 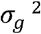. is the genetic variance, and 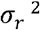 is the residual or environmental variance 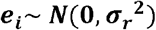.

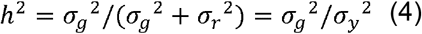

### Network Propagation using Random Walk with Restart Using Gene Centric Scores

In this study, we applied the HotNet2 algorithm to conduct network propagation analyses on the reference human PPI network (29). HotNet2 employs a random walk with restart approach, in which the restart probability is governed by the parameter R, as shown in equation 5. Each node i in the network is assigned a vector ***s***_***i***_, and the elements of the transition matrix are given by :

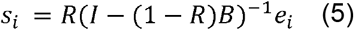

in the given context, *e*_*i*_ denotes a vector in which the i-th element is assigned a value of 1, and all other elements are set to 0. This type of vector is often referred to as a one-hot vector or a unit vector. The role of *e*_*i*_ is to specify the initial node from which the random walk process begins. The j-th element of the resulting vector s indicates the likelihood that a random walk starting from node i will terminate at node j after a predetermined number of steps or when the convergence condition is satisfied. To execute the HotNet2 algorithm, we inputted the heat scores for each gene, defined as −log (p-value) of each gene’s heritability as calculated by LDAK, into the software as the initial scores for network propagation.

### Heat Diffusion and Module Detection in Network Propagation to Uncover Functionally Coherent Subnetworks

To determine the heat for each node, the negative logarithm of the p-value, derived from the REML LDAK heritability on SNPs within 50 kilobases of the canonical gene regions, is calculated. This heat value serves as a measure of the statistical significance or importance of each node in the network. To identify statistically significant submodules within the network, edge swapping permutation tests are performed. In this study, the analysis was conducted using 500 permutations, which resulted in the discovery of 14 distinct modules that exhibited significant statistical properties.

### Heritability Partitioning Using LDAK: Module-Specific Kinship Matrix Calculation and REML Analysis

To partition heritability among different gene modules, we employed the LDAK software. The process involved calculating kinship matrices for each gene module and subsequently estimating the heritability associated with each module.

Initially, we used the LDAK command-line tool to calculate direct kinship matrices for each of the 14 predefined gene modules. The kinship matrices were calculated using the --calc-kins-direct option, with a power parameter set to -0.25, which downweighs the contribution of SNPs with higher MAFs. The -ignore-weights YES option was applied to ensure that all SNPs contributed equally unless specified otherwise. Each module-specific list of SNPs was provided through the --extract option, and the genotype data was specified using the --bfile option, pointing to the PLINK binary file set.

Following the creation of kinship matrices, heritability partitioning was conducted using LDAK’s REML (Restricted Maximum Likelihood) analysis. We specified covariates, including the first principal component and gender, through the --covar option to account for potential confounding factors. The --mgrm option was used to input a file (mgrm.list) containing the paths to the kinship matrices for all modules. The phenotype data, including the trait of interest, was provided via the --pheno option, and the specific phenotype to be analyzed was indicated using the --mpheno option.

### Imputation of Gene Expression Using PrediXcan for Tissue-Specific Analysis of Discriminatory Gene Networks

In our study, we employed the PrediXcan method to impute gene expression levels from genotype data, focusing on tissue-specific models relevant to rheumatoid arthritis (RA) endotypes. PrediXcan uses a linear model to predict gene expression based on the relationship 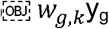 is the expression trait for gene g, and *X*_*k*_ is the number of reference allele for marker k and *w*_*g,k*_ the contribution of each *X*_*k*_ to the expression trait. This model allows for the imputation of gene expression levels purely based on genetic variation.

For our analysis, we imputed gene expression data across various tissue types, including whole blood and subcutaneous adipose tissue. These tissues were selected due to their relevance in RA pathogenesis. By applying PrediXcan, we were able to assess whether genes within specific network modules displayed distinct expression profiles across these tissue types, enabling us to identify gene networks that may be discriminatory between different RA endotypes.

After the imputation, we performed linear regression analyses to evaluate the association between imputed gene expression and the phenotypic trait of interest. This was done by merging the imputed expression data with phenotype data and fitting linear models (lm function from base r package) to examine the relationship between gene expression and two RA endotypes. The use of a linear model allowed us to quantify the performance of tissue specific network gene modules associated with RA endotypes. We use -log0(p-value) of the regression model to quantify the significance of the modules in tissue-specific discrimination between CCP-positive and CCP-negative RA.

### Evaluation of Gene Module Discriminatory Performance using Bulk RNA-seq Data

Bulk RNA-seq data and metadata from AMP RA phase 1 study were retrieved from Immport (study accession code SDY998). Bulk RNA-seq data corresponded to sorted T cells, B cells, monocytes, and fibroblasts from disaggregated synovial tissues. The methods for data collection are described in the source paper (31). CCP and RF positivity status was available for 29 patients. Of these, 19 patients were CCP+/RF+ and 10 were either singly positive (CCP-/RF+ or CCP+/RF-) or double negative (CCP-/RF-). For each cell type, we removed genes with variability below the 25th quantile for the entire cell dataset.

We assessed the discriminatory power of gene modules identified in prior network-based analyses using logistic regression with *l*_*1*_ regularization. The analysis was performed using R with several packages to facilitate data handling, model training, and parallel computation.

Specifically, the **BiomaRt** package was used for mapping Ensembl gene identifiers to gene names. Parallel processing was enabled using **doParallel** and **foreach** to expedite computations.To evaluate the predictive power of the gene modules, we employed the caret package, which is designed for training and evaluating machine learning models. Specifically, we used a logistic regression model with LASSO regularization (glmnet method in **caret**) to identify the most relevant features (genes) within each module. The model was trained using leave-one-out cross-validation, which provides a robust estimate of model performance for small datasets. The custom train control function, incorporating area under the ROC curve (AUC) as the performance metric, was implemented to select the optimal model parameters (lambda and alpha).

### Replication GWAS Using All of Us Cohort

As a replication cohort, we analyzed 248 participants from the All of Us Research Program (23). Participants were filtered to include only Caucasian individuals who were RF+ (RF > 20 IU/mL). These individuals were further stratified based on CCP levels into the two endotypes: CCP+/RF+ (n = 93) and CCP−/RF+ (n = 155). This study was approved by the All of Us Institutional Review Board.

Phenotypic data for each subject was represented by a vector **Y** with 248 entries, where a value of 1 corresponded to CCP+/RF+ status and 0 to CCP−/RF+ status. Genotype data for each individual, covering 6,879,629 SNPs, was organized into a 248 × 6,879,629 matrix **X**—each row represented an individual’s genotype and each column corresponded to a specific SNP. Quality control, including a genotype missingness filter of 10% (−-geno 0.1) to exclude SNPs with a high proportion of missing data and a MAF threshold of 5% (−-maf 0.05) to exclude rare variants, was conducted using PLINK.

### Evaluation of Gene Module Performance in CTAP and Treatment Response Classification Using scRNA-seq Data

To evaluate the predictive capacity of the gene modules in other functional contexts, we implemented a logistic regression modeling framework using scRNA-seq data from 70 RA patients (33). The dataset included expression matrices for multiple immune and stromal cell types, as well as metadata annotated with either CTAP labels or clinical treatment response categories.

For each cell type, we first generated pseudobulk expression profiles by aggregating single-cell counts across cells from the same patient. We then constructed binary classification models using genes from the modules to distinguish a given CTAP or treatment response category from all others using a one-vs-rest approach. Model training was conducted using L1-penalized logistic regression (LASSO) via the **glmnet** package in R. Five-fold cross-validation was used for CTAP classification tasks, while a 10-fold cross-validation was applied for treatment response prediction. All modeling procedures were orchestrated using the **caret** package to streamline training, hyperparameter tuning, and performance evaluation. ROC curves were computed using the **pROC** package, and AUC values were used as the primary performance metric to assess the discriminatory power of each module across CTAPs, treatment categories, and cell types. Gene expression filtering and data wrangling were performed using base R and the **dplyr** package.

## Data Availability

The individual genotype data is not available to the public. Summary statistics and validation data are available upon request.

## Code and Data Availability

All data and code associated with the manuscript are available at https://github.com/jishnu-lab/RAEndo.

## Acknowledgements

J.D. was supported in part by NIAID DP2AI164325 and a pilot grant from the Pittsburgh Autoimmunity Center for Excellence in Rheumatology. The authors acknowledge support from the University of Pittsburgh Center for Research Computing through the high-performance computing resources provided. The authors also gratefully acknowledge All of Us participants for their contributions, without whom this research would not have been possible.

