## Supplementary figures and images for "Multi-modal integration of protein interactomes with genomic and molecular data discovers distinct RA endotypes"

### Supplementary Figures

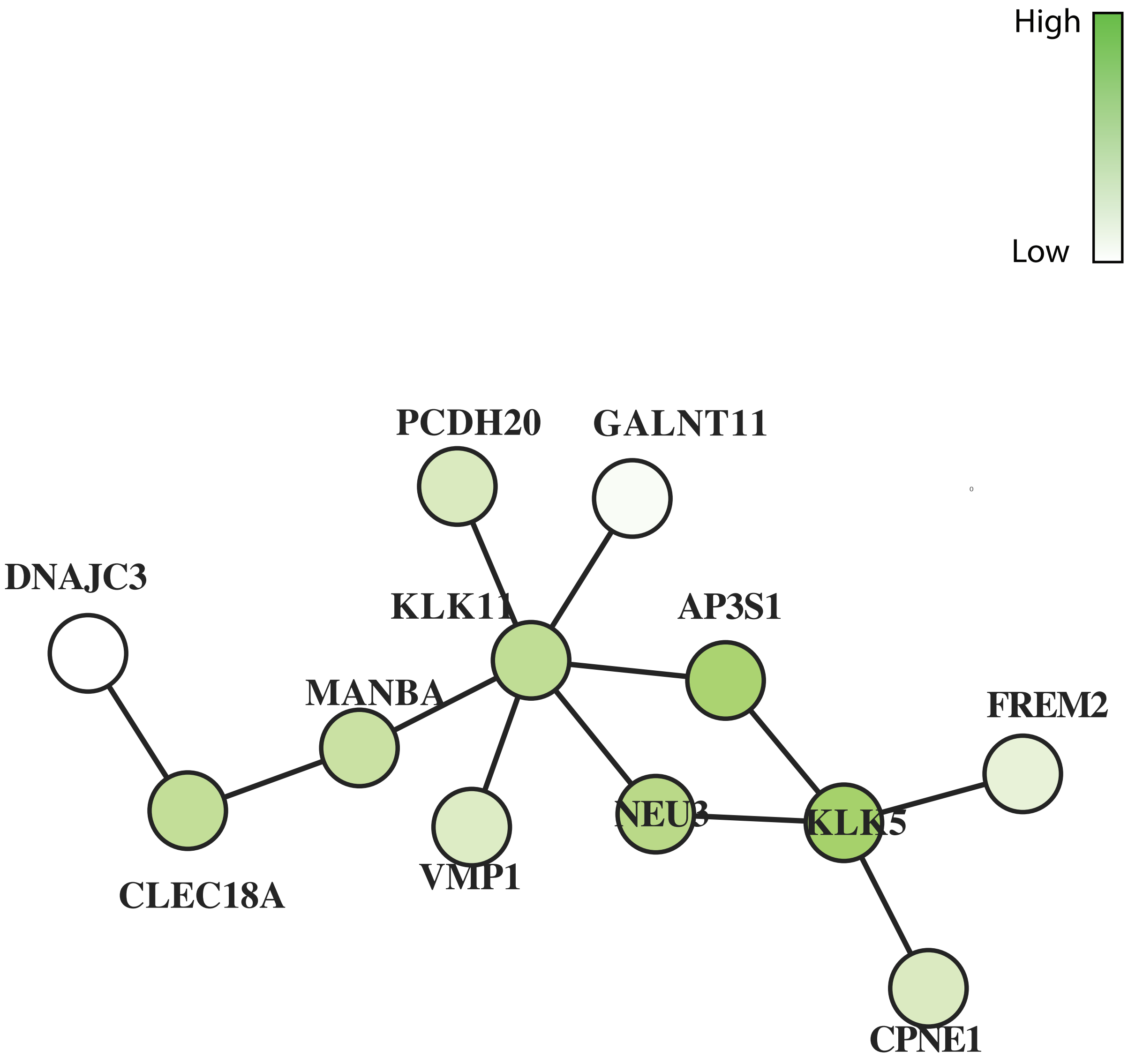
