## Supplementary Tables for "Multi-modal integration of protein interactomes with genomic and molecular data discovers distinct RA endotypes"

| Module Number | Genes in the Module | Module Name (GeneSetAI) |
|---|---|---|
| 1 | CENPBD1, MSC, ALDH1A2, PLAT, INS, IFNA2, HLA-DRA, HLA-DRB1, FGA, FGB, FGG, ALDH2, HLA-DOA, HLA-DOB, HLA-DMA, PBX2, CAMP, EPHB4, MAB21L1, STIM1, HLA-DRB5, MLKL, ORAI1, SRA1, ADAMTS1, KLK4, IFNA13, PGLYRP1, IFNA5, IFNA14, DCN, HOXB7, HLA-DMB, IFNA8, ZNF616, PHOSPHO1, GLG1, OMA1, PLA2G12B, ACAN, SH3BP1, IFNA17, IFNA6, IFNA4, SURF1, IFI44L, UBR3, SLC25A16, CHST12, ALDH6A1, LONP2, ZSCAN5A, CHTF18, TOR3A | Interferon Signaling and Extracellular Matrix Remodeling |
| 2 | SGSM2, NTN1, FSTL3, INHBA, BMP2, BMP6, ACVR2A, ACVRL1, GDF5, BMPR2, RGMB, INHBB, ENG, AFP, FNBP1L, RGMA, GDPD2, DRAXIN, ACVR1B | TGF-beta/BMP Signaling in Developmental Processe |
| 3 | HLA-DPB1, ADK, CDNF, ALKBH2, SRD5A3, NUDT15, NFS1, CLPSL1, LCN15, NDUFAF7, SUGCT, NHLRC3, SLC30A5, GCNT2, VSTM2A, ST3GAL5 | |
| 4 | RFXAP, HOXC11, IL1A, IL1B, IL1R1, IL1R2, MC4R, RFX5, STAT4, IRF4, LBH, AGRP, CIITA, TBX21, IL1RAP, PROX1 | Immune Response and Inflammation Modulation |
| 5 | GYPA, C9, C8A, C8B, C7, GOLT1A, LHFPL3, TMEM154, ANKRD29, EVA1A, C6, CPT1B, RNF24 | Complement System Activation and Regulation |
| 6 | DNAJC3, KLK5, CLEC18A, MANBA, FREM2, PCDH20, GALNT11, AP3S1, VMP1, CPNE1, KLK11, NEU3 | |
| 7 | IL4, IL2RB, PTPRD, IL2RG, IL13, IL15, IL13RA1, NAGS, LRFN5, SLITRK1, IL1RAPL1, TMEM39B | Cytokine Signaling and Immune Response Modulation |
| 8 | ENDOD1, TFF1, FGF2, FGFR2, VSNL1, ZNF695, FGF7, FGF8, PDLIM2, SRL, TEX13B, FGFBP1 | Fibroblast Growth Factor Signaling and Epithelial-Mesenchymal Interactions |

| | | |
|---|---|---|
| 9 | AKR1C4, AKR1D1, NUDT6, AKR1C1, UBR2, AKR1B15, TEX33, AKR1B10, PLXDC2, LRRC42, PDP2 | Steroid Hormone Metabolism and Detoxification |
| 10 | INS-IGF2, CANT1, OXSM, OBP2A, ENTPD6, CLN5, ERLEC1, ATF6B, OBP2B | Protein Folding and Glycosylation in the Endoplasmic Reticulum |
| 11 | HCRT, HLA-DQA1, HLA-DQB1, GALT, HCRTR2, ATP6V0A2 | |
| 12 | TIMP3, COLEC11, ADAMTS5, COL6A1, RTBDN | Extracellular matrix remodeling and tissue development |
| 13 | CCL3, CCR1, CCR3, CCL15 | Chemokine-mediated immune cell trafficking |
| 14 | CGA, FSHB, FSHR, CGB7 | Gonadotropin Synthesis and Follicle Stimulation |
